# Effects of a Cardiac and Psychosocial Rehabilitation Program on Anxiety, Depression, Self-Efficacy, Quality of Life, and Exercise Capacity in Young Adults With Fontan Circulation: A Randomized Controlled Trial

**DOI:** 10.1101/2024.09.16.24313780

**Authors:** Ju Ryoung Moon, June Huh, Jinyoung Song, I-Seok Kang, Goeun Park, Kyunga Kim, Jidong Sung, Sung-A Chang, Seung Woo Park, Ji-Hyuk Yang, Tae-Gook Jun

## Abstract

**BACKGROUND:** Young adults with Fontan circulation (FC) face psychosocial, physical, and environmental challenges and are vulnerable to emotional distress. They need holistic intervention programs to face these problems and enhance their quality of life. This study developed and evaluated a cardiac and psychosocial rehabilitation (CPR) program for young adults with FC.

**METHODS AND RESULTS:** Thirty-eight young adults with FC aged 18–23 years were prospectively recruited from an outpatient clinic and randomized into a CPR group (n = 12) that underwent eight weekly group-based sessions combining a psychosocial program based on cognitive behavioral therapy with home-based cardiac rehabilitation (CR), a CR group (n = 12) that received only home-based CR, and a usual care control group (n = 12). The study assessed CPR program effectiveness based on the Hospital Anxiety and Depression Scale, self-efficacy, quality of life, and exercise capacity testing at three time points: pre-intervention, post-intervention, and four weeks post-intervention. Participants’ average ages were 19.75 (2.28), 19.65 (1.79), and 20.38 (1.85) years in the CPR, CR, and control groups, respectively; 50% (CPR group) to 53.8% (CR and control groups) of the participants were male. After eight weeks, depression and anxiety decreased, whereas self-efficacy and quality of life increased in the CPR group compared with the CR and control groups. The groups showed no differences in exercise capacity.

**CONCLUSION:** CPR can be part of an intervention for alleviating psychological distress and improving self-efficacy in young adults with FC. Efforts should be made to increase the efficacy of home-based CR.

## INTRODUCTION

Fontan palliation is the last of several surgeries used to treat single-ventricle congenital heart disease (CHD), a class of severe complex CHD in which two-ventricle circulation cannot be established.^1^ Owing to advances in diagnosis and treatment, the 30-year survival rate of patients who undergo Fontan circulation (FC) currently exceeds 80%.^2,3^ The number of adults with FC is rapidly increasing. An estimated 70,000 adults are living with FC globally, and this number is expected to double over the next 20 years.^4^ However, patients who undergo FC may experience progressive functional limitations, serious cardiac diseases, non-cardiac morbidity, and the possibility of Fontan circulatory failure, which may be severe enough to require heart transplantation or cause premature death.^1^ Thus, patients with FC experience psychological and physical difficulties throughout their lives.^5^

The lifetime prevalence of mental disorders such as depression and anxiety in patients with FC is estimated at 65%, much higher than the 11% prevalence in healthy peers.^5,6^ A meta-analysis of psychological function in patients with single-ventricle CHD identified 24 studies that evaluated the risk of internalizing conditions such as depression and anxiety.^7^ Additionally, MRI studies of adolescents’ brains with single-ventricle CHD have reported damage to select areas that control depression and anxiety, providing evidence for the functional deficits observed.^8,9^ Therefore, management guidelines for patients with FC should include interventions that address psychosocial issues such as depression and anxiety.^3,9,10^

Cognitive behavioral therapy (CBT) is a non-pharmacological psychological intervention focused on challenging and modifying maladaptive cognitions and behaviors.^11^ CBT has a strong evidence base supporting its use in the treatment of depression and anxiety in both children^12,13^ and adults, including in primary care settings and adults with acquired cardiovascular disease.^14–19^ However, studies on CBT-based interventions in adults with CHD are scarce. Kovacs et al. conducted a pilot study evaluating the feasibility of group-based CBT (eight 90-minute sessions) in adults with heterogeneous CHD diagnoses.^20^ They found a moderate effect size in favor of the therapy for depressive symptoms, although the sample size was small.^20^ Nonetheless, their findings can serve as a basis for applying CBT-based interventions to reduce psychological problems and improve quality of life for adults with CHD. However, no study has explored its application to patients with FC experiencing depression and anxiety.^9,12,21,22^

Cardiac rehabilitation (CR), exercise, and exercise training are non-pharmacological psychological interventions that can be applied in addition to CBT for adolescents and adults.^12^ Historically, exercise participation in patients with FC is not recommended owing to risks associated with their complex cardiac physiology, including hemodynamic derangement and sudden cardiac death.^23^ However, reduced exercise capacity in this cohort has also been associated with a worse long-term prognosis.^24,25^ Given the impaired exercise tolerance of most patients with FC, health care teams should encourage appropriate physical activity and dispel myths about activity limitations.^26^

The relation between exercise training and both improved mental health and overall quality of life (QOL) is well-established and widely applied clinically and in healthy populations.^27–29^ Participating in physical activity improves psychosocial functioning by increasing opportunities to socialize and connect with peers.^30^ In patients with FC, exercise training is associated with improvements in not only physical functioning but also psychosocial functioning and QOL. However, although exercise programs can safely and effectively improve the health and well-being of patients with FC, the results are somewhat inconsistent.^31,32^

Therefore, we sought to confirm the effectiveness of a program combining a CBT-based psychological intervention and CR in improving psychosocial function and QOL among young adults with FC and symptoms of depression and anxiety.

## METHODS

### Study Design and Patients

This study was an open-label, prospective, randomized controlled trial (RCT).^33^ Participants were randomly assigned to three groups: a cardiac and psychosocial rehabilitation (CPR) group receiving CBT-based psychological rehabilitation (CPR) and CR; a CR group receiving only CR; and a control group receiving usual care. Five intervention outcomes— anxiety, depression, self-efficacy, QOL scores, and exercise test results—were assessed pre-intervention, immediately post-intervention, and at a four-week follow-up (Figure 1). The study targeted young adults who had undergone FC and were registered at the Samsung Medical Center.

**Figure 1.**
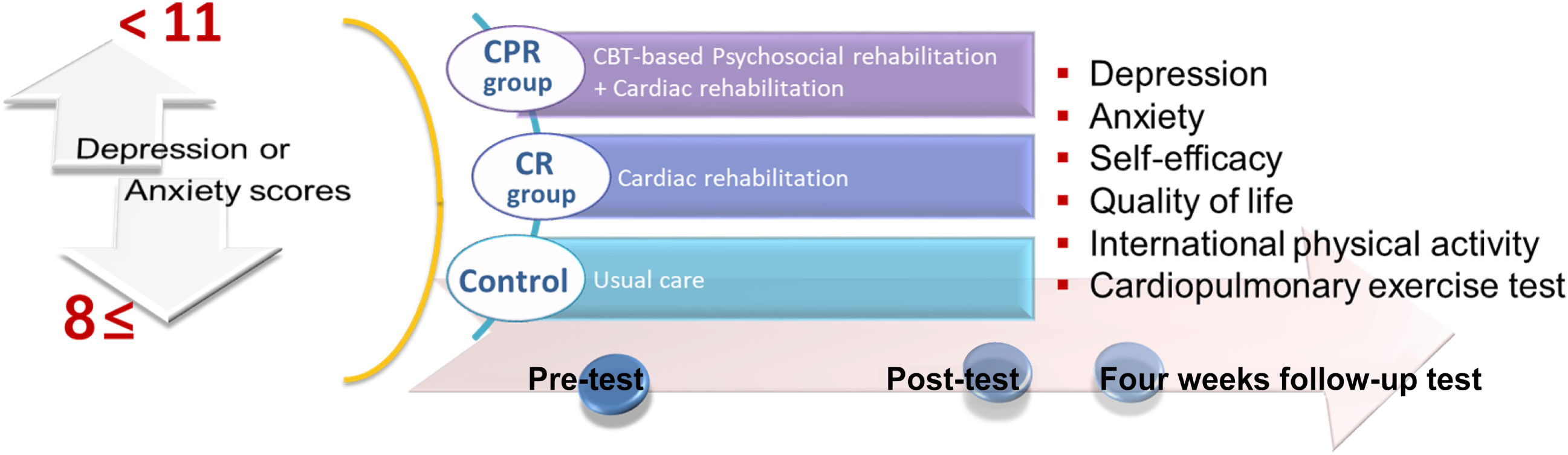
Procedure and outcome assessment. CPR indicates, cardiac and psychosocial rehabilitation; and CR, cardiac rehabilitation.

We measured psychological distress using the Hospital Anxiety and Depression Scale (HADS).^34^ The specific enrollment and exclusion criteria for participants were as follows:

*Inclusion criteria:*

--Aged 18 to 23 years more than one year after Fontan surgery
- Had no surgery planned during study participation
- Had scores ≥ 8 for the anxiety (HADS-A) and/or depression (HADS-D) HADS subscales^19,35^
- Could read and complete the consent form and questionnaire, and participate in a Korean-language group

*Exclusion criteria:*

- Undergoing psychotherapy during study participation
- Had significant cognitive impairment, psychosis, or personality disorder noted in their medical record
- Had scores ≥ 11 for HADS-D and > 17 for the Beck Depression Inventory (BDI) as evaluated by a psychiatrist or had been identified with severe depression or other serious psychiatric diagnoses^12,19^
- Exhibited indications for the restriction of exercise of greater-than-moderate intensity based on recommendations from the American Heart Association^36,37^

The study was approved by the Institutional Review Board of the Samsung Medical Center and prospectively registered with the Korea Clinical Trials Registry (cris.nih.go.kr, KCT0007737).

The study’s sample size was calculated using PASS 2022 (version 22.0.2, NCSS, LLC; Kaysville, Utah, USA). A sample size of 12 in each group was required to detect mean differences in the depression change score of 2.02 with a standard deviation of 1.28,^20^ using a two-sided two-samples *t*-test with a power of 80% and a significance level of 2.5% for the first co-primary comparison and 1.25% for the second.

### Interventions

Before the study began, we prepared a booklet containing the goals of the investigation, program contents, and worksheets for the activities to be completed by the participants. The study’s intervention programs were CPR and CR. In the CPR program, we provided a CPR intervention aimed at helping patients cope with their CHD, manage distress (depression and anxiety), and improve their social coping skills. The foundations and accepted learning theories of CBT served as the basis for the manualization of the therapeutic procedures.^38,39^ The CPR program had eight sessions divided into three stages: “knowing my heart,” “cognitive restructuring and alleviating distress symptoms,” and “promoting social coping skills” (Table 1). Every session included homework, and the participants were given the opportunity to offer comments at the conclusion of each session. The group CPR program ran for eight weeks, with weekly meetings of 90 minutes each. Three subgroups, each consisting of three to four participants,^19^ finished the eight-week CPR program. The program was held at a time convenient for the participants (e.g., weekday evenings and Saturday mornings) in a cozy, roomy counseling room at the hospital. The sessions were led by a psychologist with over 20 years of experience performing CBT-based programs and group interventions for patients with mental disorders in a tertiary center’s psychiatry department.

**Table 1.**
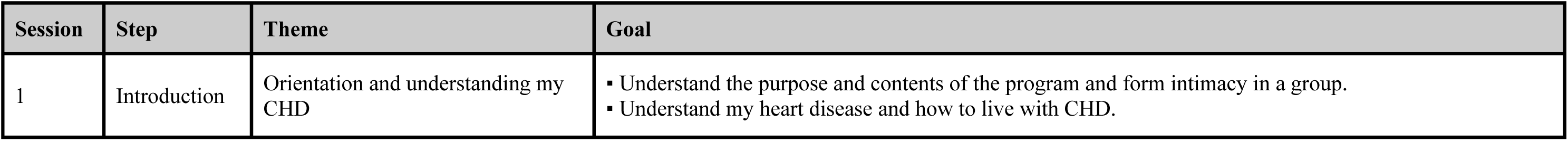
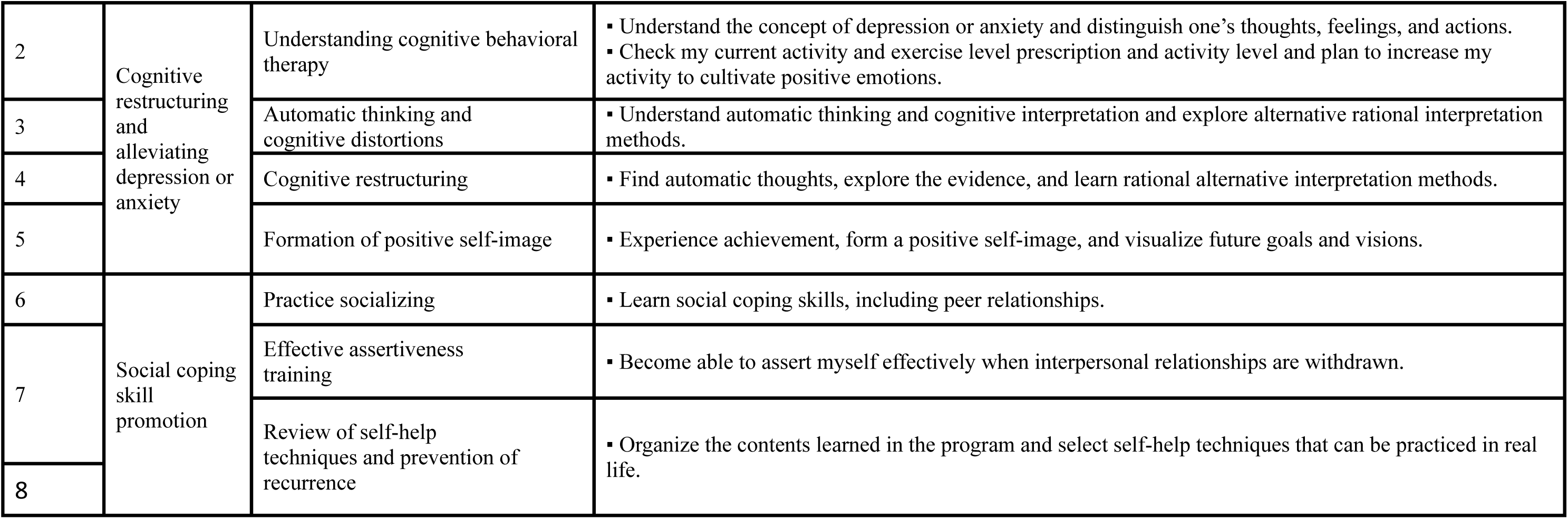
Contents of the Eight Sessions of the CBT-Based Psychosocial Rehabilitation. CBT indicates cognitive behavioral therapy; and CHD, congenital heart disease.

| Session | Step | Theme | Goal |
| --- | --- | --- | --- |
| 1 | Introduction | Orientation and understanding my CHD | ▪ Understand the purpose and contents of the program and form intimacy in a group.<br>▪ Understand my heart disease and how to live with CHD. |
| 2 | Cognitive restructuring and alleviating depression or anxiety | Understanding cognitive behavioral therapy | ▪ Understand the concept of depression or anxiety and distinguish one's thoughts, feelings, and actions.<br>▪ Check my current activity and exercise level prescription and activity level and plan to increase my activity to cultivate positive emotions. |
| 3 |  | Automatic thinking and cognitive distortions | ▪ Understand automatic thinking and cognitive interpretation and explore alternative rational interpretation methods. |
| 4 |  | Cognitive restructuring | ▪ Find automatic thoughts, explore the evidence, and learn rational alternative interpretation methods. |
| 5 |  | Formation of positive self-image | ▪ Experience achievement, form a positive self-image, and visualize future goals and visions. |
| 6 | Social coping skill promotion | Practice socializing | ▪ Learn social coping skills, including peer relationships. |
| 7 |  | Effective assertiveness training | ▪ Become able to assert myself effectively when interpersonal relationships are withdrawn. |
| 8 |  | Review of self-help techniques and prevention of recurrence | ▪ Organize the contents learned in the program and select self-help techniques that can be practiced in real life. |

We gave the therapist a standardized manual and described the recruitment training process to ensure consistency among the treatments in the study.^39^ The therapist adhered to the CBT guidebook.^39^ Two CBT specialists who were blind to the treatment condition monitored the therapist’s adherence. They used audio recordings and written materials (e.g., homework assignments, protocol session checklists) to rate adherence to the treatment manual’s modules and treatments and compliance with CBT’s guiding principles.

The study’s CR program was an eight-week home-based moderate-to-high-intensity aerobic exercise program.^40–42^ Patients received one-on-one training sessions with a physiotherapist at the rehabilitation site on the pre-test evaluation day. During this session, the participants received instructions on the components of the program, an individualized exercise program based on cardiopulmonary exercise testing (CPET) results at the pre-test, and symptom monitoring methods based on the rate of perceived exertion during exercise.^43^ The patients were also asked to complete a home exercise diary to monitor compliance and exercise quantity.

The program’s exercise intensity was approximately 60% to 70% of the participant’s maximum heart rate.^44^ Home-based exercise began at a rate of at least three times a week and was increased to a goal rate of five to seven times a week. The time started at 15–30 minutes per session and increased to 40–60 minutes per session. Depending on each patient’s functional status, the activity level started at 3,000–5,000 steps per day and increased to a goal rate of 7,000–10,000 steps per day over 2–3 weeks. When patients returned to the hospital’s rehabilitation facility at week four or five of the program, the same physiotherapist evaluated them for compliance and progress. Workout adjustments were made during the visit to improve compliance and increase exercise quantity. The onset of symptoms and fatigue after exercise was monitored over the telephone for eight weeks by a highly skilled cardiac nurse. The nurse verified that the patients were complying and modifying or progressing their exercises while ensuring their safety. From the first to the fourth week, the nurse conducted one weekly phone monitoring session; from the fifth to the eighth week, two weekly phone monitoring sessions were conducted The CPR group underwent eight sessions of the group CPR program and the CR program.

The CR group was offered only the CR program. Patients who wanted to undergo the CPR program were given one after the completion of the study. Meanwhile, the control group received no interventions during the study period (i.e., only usual care). Patients who wanted to receive the CPR and CR programs underwent these after the study was completed (Figure 1).

### Outcomes

#### Primary outcome

The primary outcome of the study was depression. We assessed depression using the HADS,^34^ which is composed of two subscales: the depression subscale (HADS-D), with even-numbered items measuring seven depressive symptoms, and the anxiety subscale (HADS-A), featuring odd-numbered items assessing seven anxiety symptoms. Each item is scored on a scale from 0 to 3, contributing to a total possible score ranging from 0 to 21. Higher scores indicate a greater level of depression or anxiety symptoms. Patients are classified into three groups on each subscale: (a) no symptoms (0–7 points), (b) mild symptoms (8–10 points), and (c) moderate or severe symptoms (≥ 11 points).^34^ This grouping represents the optimal sensitivity of both HADS-D and HADS-A in identifying clinical cases.^35^

The psychiatrist conducted assessments for patients with a HADS-D score of ≥ 11 and a BDI score of > 17. Following clinical evaluation, individuals with severe depression or other significant psychiatric diagnoses were excluded from the study. Patients who declined this assessment or were recommended for alternative treatments post-psychiatric evaluation were also excluded.^19^

#### Secondary outcomes

We assessed anxiety using the HADS-A.^34^ We measured self-efficacy using the Korean version of the General Self-Efficacy Scale (GSE),^45^ which was developed by Schwarzer and Jerusalem and then translated to and validated in Korean by Schwarzer et al.^46^ The GSE consists of 10 questions, each scored on a scale from 1 to 4, contributing to a total possible score ranging from 10 to 40. Higher scores indicate higher perceived self-efficacy. The GSE had sufficient internal consistency (Cronbach’s α > .80) and test-retest reliability (*r* = .57– .71).^45^ For this study, the internal consistency was .89.

We evaluated QOL using the Linear Analog Scale (LAS), consisting of a 10-centimeter line that is vertically oriented and graded with indicators ranging from 0 (the lowest possible QOL) to 100 (the highest possible QOL). Patients were asked to identify on a scale how good or poor their overall QOL was by marking the corresponding point. The validity, reliability, and responsiveness of the LAS for its intended application among adults with CHD have been demonstrated through its psychometric qualities.^47^

We used the International Physical Activity Questionnaire (IPAQ) short form to assess physical activity (PA) levels over the past seven days.^48,49^ PA can be calculated in two ways using the IPAQ response results of categorical and continuous variables. Categorical variables include vigorous, moderate, or walking activities. The questionnaire classified the activity levels into three categories: health-enhanced PA (HEPA), minimal activity, and inactivity.^50^

The PA for the continuous variables was calculated as the metabolic equivalent of task minutes per week (MET-min/week), according to the IPAQ analysis guide. We calculated METs by summing the total time and intensity of participation in activities in the three categories. The work intensity and each category of the vigorous- and moderate-intensity MET values were calculated as 8.0 and 4.0, respectively. Higher MET-min/week scores indicated higher PA levels. Finally, we calculated the PA levels into the MET as “Time spent in PA” × “number of times per week” × “MET level (8 or 4 METs).”^11^ The IPAQ shows acceptable reliability and validity and has been demonstrated to be a valid measurement tool for assessing PA levels in individuals with CHD.^51,52^

All exercise tests were performed by a trained physiologist at the same time of day for each patient. Patients underwent progressive symptom-limited cardiopulmonary exercise testing on a treadmill (Quinton-TM55, Quinton Cardiology Inc., Bothell, WA, USA) according to the Modified Bruce Protocol.^53,54^ Electrocardiographic monitoring and breath-by-breath exhaled gas analysis were performed using the Medical Graphics exercise testing system (Carefusion Vmax Encore 29, Yorb Linda, CA,USA).

The cardiopulmonary exercise test measured maximal oxygen uptake (VO_2_ max), and the exercise stress test measured METs and exercise time.^55^

### Randomization

We employed a randomized permuted block design with a block size of six^33^ for each of the four strata using R 3.5.2 to randomly allocate eligible patients to the CPR, CR, and control groups at a 1:1:1 allocation ratio. Stratification was performed according to sex and the New York Heart Association’s functional classification.^21,56,57^ The study director stored randomly generated assignment sequences in sealed opaque envelopes until group allocation was performed. A research assistant, who was not a researcher in this study, divided the patients into groups according to the sequence of their entry into the study by opening each envelope sequentially. The patients were divided into the CPR, CR, and control groups according to the distribution sequence written on the envelope.

After all baseline evaluations were completed, patients were notified of their assignments. The interventionists and patients could not be blinded to the interventions because of the intervention’s nature. Patients were instructed not to reveal their group assignments to the qualified assessors during the evaluations. Additionally, the patient group codes were assigned before statistical analysis, and the biostatistician was not informed of the codes’ definitions.

### Statistical analysis

We conducted statistical analyses using SAS, version 9.4 (SAS Institute Inc., Cary, NC, USA). Continuous variables were expressed as means ± standard deviations, whereas categorical variables were presented as numbers (percentages). Differences among the three groups were analyzed using the chi-squared test or Fisher’s exact test for the categorical variables and the analysis of variance for the continuous variables, based on the normality assumption.

To compare mean changes in depression among the groups, we conducted a linear regression analysis after adjusting for baseline values and stratification variables (sex and functional class). For primary comparisons, the statistical significance level was set at 0.025; for secondary comparisons, it was set at 0.0125. For secondary outcomes, we analyzed the differences in mean changes over time among the groups using a marginal regression model with generalized estimating equations. The model included group, time, and group-by-time interaction and stratification variables. For the working correlation matrix, we used the autoregressive (AR (1)) structure, which assumes decreasing correlations over time. The model presented the estimated mean and standard error. A two-tailed P-value of *P* < 0.05 was considered statistically significant.

## RESULTS

### Participants

Of the 83 patients assessed for eligibility, 18 declined to participate in the study, and 10 were excluded because they did not meet the inclusion criteria. Furthermore, 13 patients with HADS-D and HADS-A scores < 8 were also excluded. This left 41 patients with HADS-D or HADS-A scores ≥ 8. Among them, four patients with HADS-D scores ≥ 11 and BDI scores > 17 were excluded based on the psychiatrist’s recommendation for another treatment. Using a computer program, we randomly selected 38 eligible patients, with 13 allocated to the CPR group, 13 to the CR group, and 12 to the control group. One patient in the CPR group withdrew consent to participate immediately after the second session because of the long distance from their home to the hospital, and one patient in the CR group refused to take the post-test for personal reasons. This left 36 patients as study participants (Figure 2).

**Figure 2.**
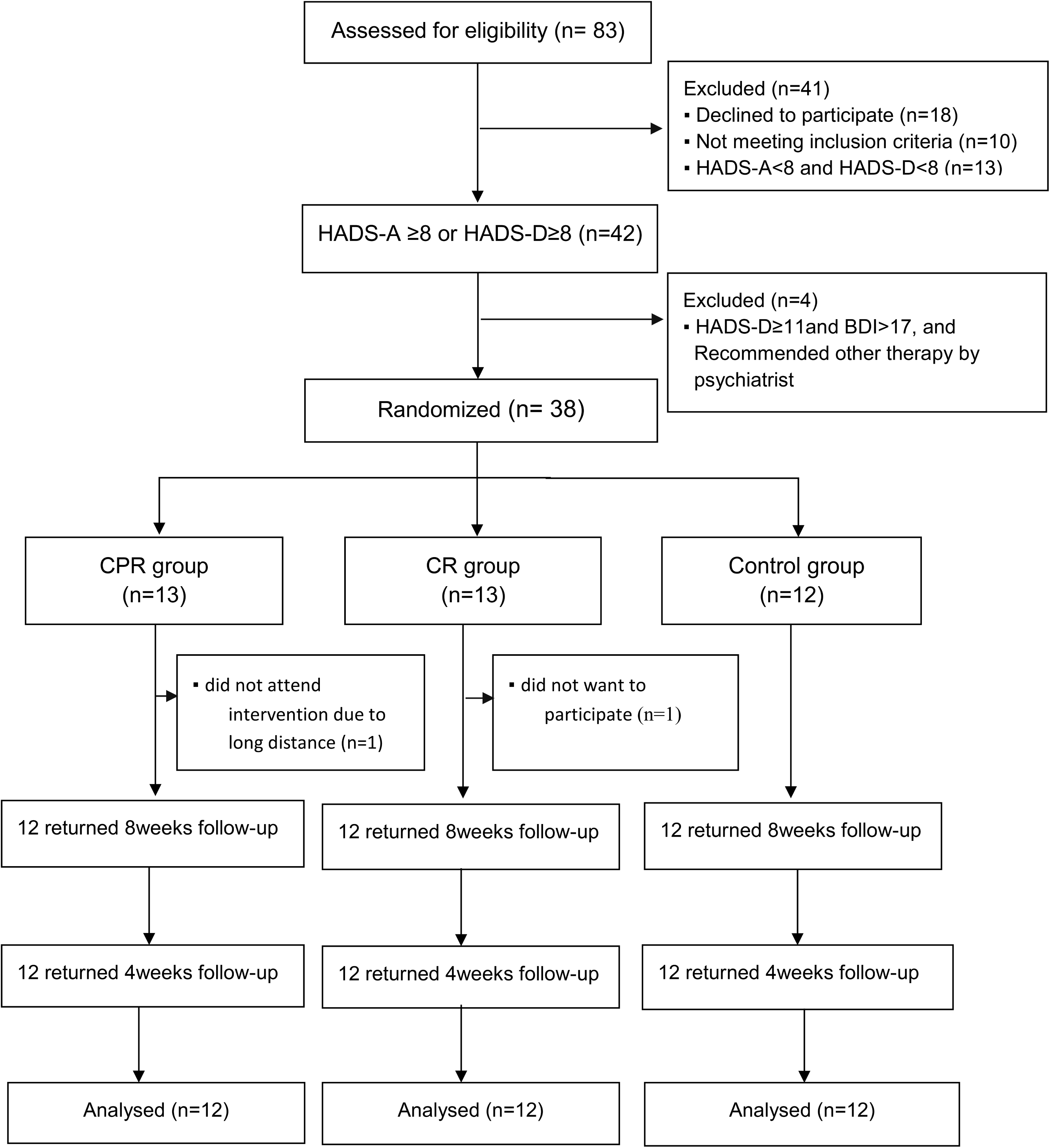
Flow diagram of progress through the phases of a randomized trial in the three groups. HADS-A indicates hospital anxiety and depression scale-anxiety; HADS-D, hospital anxiety and depression scale-depression; BDI, beck depression inventory; CPR, cardiac and psychosocial rehabilitation; and CR, cardiac rehabilitation.

Table 2 presents the demographic and clinical summary data for the three groups. The patients’ mean age was 19.83 years (1.85), with 50% being males. The CPR, CR, and control groups had similar demographic and clinical characteristics, except that the CPR group had a significantly higher percentage of patients with a dominant right ventricle (58.3% vs. 16.6% vs. 8.3%, *P* = 0.013; Table 2). We found no significant differences among the three groups in baseline scores for depression, anxiety, and secondary outcomes.

**Table 2.** Demographic and Clinical Characteristics of the Three Groups at Baseline.

|  | **Total**<br>**(N = 36)** | **CPR group**<br>**(N = 12)** | **CR group**<br>**(N = 12)** | **Control group**<br>**(N = 12)** | **P-value** |
| --- | --- | --- | --- | --- | --- |
| Age (years) | 19.83 ± 1.85 | 19.58 ± 2.19 | 19.75 ± 1.86 | 20.16 ± 1.58 | 0.742 |
| Sex, Male, n (%) | 18 (50.0) | 6 (50.0) | 6 (50.0) | 6 (50.0) | 1.000 |
| Body weight (kg) | 60.00 ± 10.55 | 59.20 ± 7.61 | 63.70 ± 14.44 | 57.11 ± 7.92 | 0.303 |
| BMI (kg/m<sup>2</sup>) | 21.67 ± 3.68 | 21.12 ± 1.79 | 22.98 ± 5.03 | 20.91 ± 3.43 | 0.328 |
| Religion, Yes, n (%) | 24 (66.6) | 8 (66.6) | 8 (66.6) | 8 (66.6) | 1.000 |
| **Education, n (%)** |  |  |  |  | 0.894 |
| High school | 17 (47.2) | 5 (41.6) | 6 (50.0) | 6 (50.0) |  |
| College | 19 (52.7) | 7 (58.3) | 6 (50.0) | 6 (50.0) |  |
| **Family type, n (%)** |  |  |  |  | 0.766 |
| Extended | 1 (2.7) | 0 (0.0) | 1 (8.3) | 0 (0.0) |  |
| Nuclear | 25 (69.4) | 9 (75.0) | 8 (66.6) | 8 (66.6) |  |
| Single-parent | 8 (22.2) | 2 (16.6) | 3 (25.0) | 3 (25.0) |  |
| Alone | 2 (5.5) | 1 (8.3) | 0 (0.0) | 1 (8.3) |  |
| **Occupation, n (%)** |  |  |  |  | 0.737 |
| White-collar | 11 (30.5) | 4 (33.3) | 4 (33.3) | 3 (25.0) |  |
| Professional | 3 (8.3) | 2 (16.6) | 0 (0.0) | 1 (8.3) |  |
| Service | 10 (27.7) | 2 (16.6) | 5 (41.6) | 3 (25.0) |  |
| Labor | 3 (8.3) | 0 (0.0) | 1 (8.3) | 2 (16.6) |  |
| Unemployed | 2 (5.5) | 1 (8.3) | 0 (0.0) | 1 (8.3) |  |
| Student | 7 (19.4) | 3 (25.0) | 2 (16.6) | 2 (16.6) |  |
| **Occupation type, n (%)** |  |  |  |  | 0.744 |
| Full time | 21 (58.3) | 7 (58.3) | 8 (66.6) | 6 (50.0) |  |
| Part time | 9 (25.0) | 2 (16.6) | 3 (25.0) | 4 (33.3) |  |
| **Monthly income, n (%)** |  |  |  |  | 0.940 |
| Low | 3 (8.3) | 1 (8.3) | 1 (8.3) | 1 (8.3) |  |
| Middle | 21 (58.3) | 6 (50.0) | 8 (66.6) | 7 (58.3) |  |
| High | 12 (33.3) | 5 (41.6) | 3 (25.0) | 4 (33.3) |  |
| **Oxygen saturation (%), room air** | 91.14 ± 3.40 | 91.00 ± 4.13 | 91.33 ± 2.74 | 91.25 ± 3.49 | 0.971 |
| **Dominant ventricle, n (%)** |  |  |  |  | 0.013 |
| Right ventricle | 10 (27.7) | 7 (58.3) | 2 (16.6) | 1 (8.3) |  |
| Left ventricle | 26 (72.2) | 5 (41.6) | 10 (83.3) | 11 (91.6) |  |
| **Heterotaxy, n (%)** |  |  |  |  | 0.522 |
| Right isomerism | 7 (19.4) | 3 (25.0) | 1 (8.3) | 3 (25.0) |  |
| Left isomerism | 1 (2.78) | 0 (0.0) | 1 (8.3) | 0 (0.0) |  |
| None | 28 (77.7) | 9 (75.0) | 10 (83.3) | 9 (75.0) |  |
| **Frequency of surgery** | 2.91 ± 1.46 | 3.25 ± 1.91 | 3.33 ± 1.30 | 2.16 ± 0.71 | 0.089 |
| **Years since Fontan** | 18.09 ± 4.07 | 17.89 ± 5.86 | 17.01 ± 2.13 | 19.38 ± 3.28 | 0.364 |
| **Fontan type, n (%)** |  |  |  |  | 0.199 |
| F-lateral Fontan | 7 (19.4) | 4 (33.3) | 0 (0.0) | 3 (25.0) |  |
| Non-F-lateral Fontan | 2 (5.5) | 1 (8.3) | 0 (0.0) | 1 (8.3) |  |
| F-ECC Fontan | 23 (63.8) | 7 (58.3) | 9 (75.0) | 7 (58.3) |  |
| Non-F-ECC Fontan | 4 (11.1) | 0 (0.0) | 3 (25.0) | 1 (8.3) |  |
| **Fenestration status, n (%)** |  |  |  |  | 0.676 |
| Spontaneous closure | 9 (25.0) | 5 (41.6) | 2 (16.6) | 2 (16.6) |  |
| Device closure | 15 (41.6) | 5 (41.7) | 5 (41.6) | 5 (41.6) |  |
| Patent | 6 (16.6) | 1 (8.3) | 2 (16.6) | 3 (25.0) |  |
| None | 6 (16.6) | 1 (8.3) | 3 (25.0) | 2 (16.6) |  |
| **NYHA class, n (%)** |  |  |  |  | 1.000 |
| Class I | 9 (25.0) | 3 (25.0) | 3 (25.0) | 3 (25.0) |  |
| Class II | 27 (75.0) | 9 (75.0) | 9 (75.0) | 9 (75.0) |  |
| **Depression** | 7.61 ± 1.20 | 7.66 ± 1.07 | 7.50 ± 1.38 | 7.66 ± 1.23 | 0.929 |
| **Anxiety** | 8.55 ± 1.36 | 8.75 ± 1.21 | 8.50 ± 1.44 | 8.41 ± 1.50 | 0.831 |
| **Quality of life** | 79.44 ± 9.42 | 79.66 ± 9.49 | 78.91 ± 9.20 | 79.75 ± 10.35 | 0.973 |
| **Self-efficacy** | 2.72 ± 0.35 | 2.75 ± 0.34 | 2.71 ± 0.36 | 2.72 ± 0.38 | 0.948 |
| **Physical activity level, n, (%)** |  |  |  |  | 0.940 |
| Inactive | 10 (27.7) | 4 (33.3) | 3 (25.0) | 3 (25.0) |  |
| Minimally active | 15 (41.6) | 4 (33.3) | 6 (50.0) | 5 (41.6) |  |
| Health enhancing physical activity | 11 (30.5) | 4 (33.3) | 3 (25.0) | 4 (33.3) |  |
| **IPAQ, physical activity (METs-min/week)** | 699.98 ± 507.14 | 697.37 ± 578.06 | 698.33 ± 482.92 | 704.20 ± 500.96 | 0.999 |
| **CPET** |  |  |  |  |  |
| Exercise time (min) | 9.96 ± 1.68 | 9.69 ± 2.08 | 9.83 ± 1.35 | 10.36 ± 1.58 | 0.606 |
| Metabolic equivalent (METs) | 8.39 ± 1.84 | 8.14 ± 2.21 | 8.34 ± 1.34 | 8.70 ± 1.99 | 0.765 |
| Maximal oxygen uptake (VO<sub>2</sub> max; mL/Kg/min) | 28.91 ± 6.58 | 28.51 ± 7.77 | 27.80 ± 5.05 | 30.40 ± 6.94 | 0.620 |

Data are expressed as mean ± standard deviation or number (percentage).
BMI indicates body mass index; F-lateral, fenestration - lateral; F-ECC, fenestration-extracardiac conduit; NYHA, New York Heart Association; IPAQ, international physical activity questionnaire; and CPET, cardiopulmonary exercise testing.

### Primary Outcomes

The results for the primary outcome variables are presented in Table 3 and Figure 3a. Depression scores declined at post-intervention and follow-up tests from the baseline in the CPR group but not in the control group. The depression scores decreased significantly post-intervention (–2.00 ± 0.42) in the CPR group compared with the control (0.25 ± 0.75, *P* for difference < 0.0001) and CR groups (–0.66 ± 1.03, *P* for difference = 0.003). These differences were maintained at the four-week follow-up, with the most significant decreases observed in the CPR group (–2.25 ± 0.62, *P* for difference < 0.0001). We also observed a substantial improvement in the CR group compared with the control group (Table 3). Depression scores declined significantly according to group (*P* = 0.005) and time (*P* < 0.001). We thus observed a significant group-by-time interaction (*P* < 0.001; Figure 3a).

**Table 3.** Depression and Anxiety: Comparison of Changes From Baseline Among Groups.

|  |  |  |  | Diff. (=Post-test (F/U test) – Pre-test) |  |  | *P*-value adjusted baseline value and stratified variables |  |  |
| --- | --- | --- | --- | --- | --- | --- | --- | --- | --- |
|  | CPR group (A) | CR group (B) | Control group (C) | CPR group | CR group | Control group | Primary comparison | Secondary comparison |  |
| Outcome | (N = 12) | (N = 12) | (N = 12) | (N = 12) | (N = 12) | (N = 12) | A vs. C | B vs. C | A vs. B |
| **Depression (scores)** |  |  |  |  |  |  |  |  |  |
| Pre-test | 7.66 ± 1.07 | 7.50 ± 1.38 | 7.66 ± 1.23 |  |  |  |  |  |  |
| Post-test | 5.66 ± 1.07 | 6.83 ± 0.83 | 7.91 ± 1.16 | –2.00 ± 0.42 | –0.66 ± 1.07 | 0.25 ± 0.75 | < 0.0001 | 0.0026 | 0.0002 |
| Follow-up test | 5.41 ± 0.79 | 7.16 ± 0.83 | 7.91 ± 0.90 | –2.25 ± 0.62 | –0.33 ± 0.77 | 0.25 ± 0.86 | < 0.0001 | 0.0085 | < 0.0001 |
| **Anxiety (scores)** |  |  |  |  |  |  |  |  |  |
| Pre-test | 8.75 ± 1.21 | 8.50 ± 1.44 | 8.47 ± 1.50 |  |  |  |  |  |  |
| Post-test | 6.75 ± 0.96 | 7.66 ± 1.43 | 8.25 ± 1.81 | –2.00 ± 0.85 | –0.83 ± 0.71 | –0.16 ± 0.83 | < 0.0001 | 0.0604 | 0.002 |
| Follow-up test | 5.83 ± 0.83 | 7.83 ± 0.83 | 8.66 ± 1.15 | –2.91 ± 0.99 | –0.66 ± 1.15 | 0.25 ± 1.05 | < 0.0001 | 0.0158 | < 0.0001 |

Data are expressed as mean ± standard deviation.
CPR indicates cardiac and psychosocial rehabilitation; and CR, cardiac rehabilitation.

**Figure 3.**
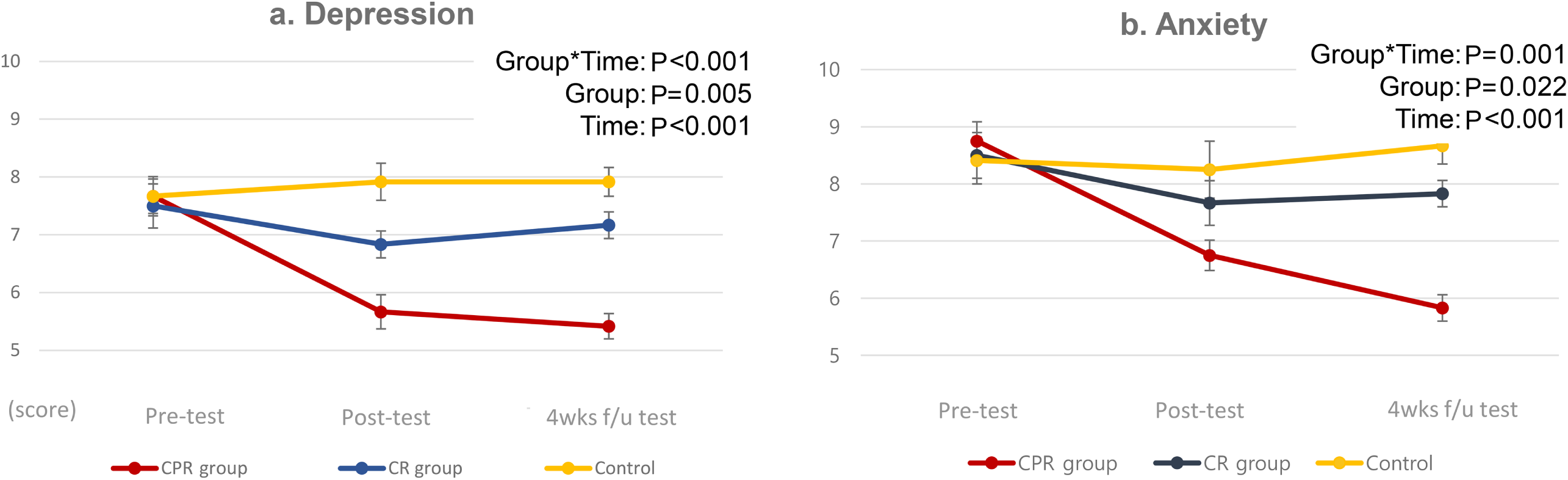
Changes in hospital depression and anxiety scale scores of the CPR, CR and control groups. Estimated score for depression (a) and anxiety (b) at pre-, post-, and four-week follow-up for the three groups from a marginal regression model with generalized estimating equations adjusted for sex and functional class.

### Secondary Outcomes

Anxiety scores declined at the post-intervention and follow-up tests from the baseline in the CPR group but not in the control group. Anxiety scores decreased significantly post-intervention (–2.00 ± 0.85) in the CPR group compared with the control (–0.17 ± 0.83, *P* for difference < 0.0001) and CR groups (–0.83 ± 0.72, *P* for difference = 0.002). These differences were maintained at the four-week follow-up, with the most significant decreases observed in the CPR group (–2.92 ± 0.99, *P* for difference < 0.0001). However, we found no substantial decrease in CR relative to the control group (Table 3). Anxiety scores declined significantly according to group (*P* = 0.022) and time (*P <* 0.001). We thus observed a significant group-by-time interaction (*P =* 0.001; Figure 3b).

Self-efficacy scores improved by 0.56 ± 0.07 in the CPR group compared with an improvement of 0.05 ± 0.07 in the CR group (*P* for difference < 0.001). These differences were maintained at follow-up (*P* for difference < 0.001). The control group showed no significant changes in the self-efficacy scores in the pre-, post-, and follow-up tests. Self-efficacy scores declined significantly according to group (*P* = 0.006) and time (*P* = 0.001), revealing a significant group-by-time interaction (*P* = 0.002; Figure 4a).

**Figure 4.**
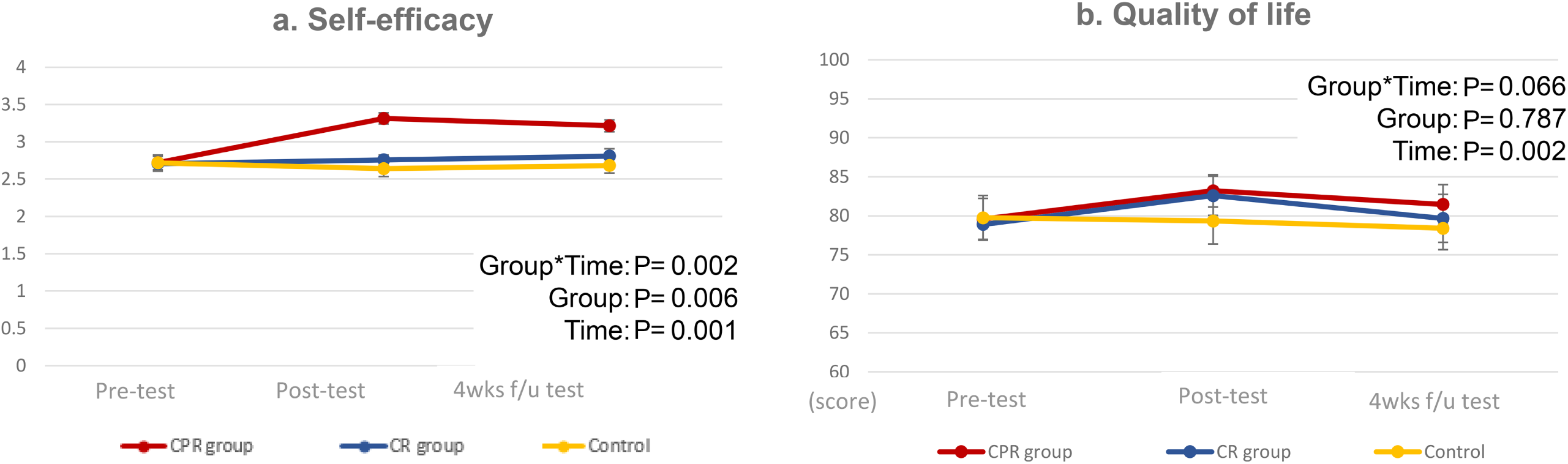
Changes in self-efficacy and quality of life in the CPR, CR and control groups. Estimated score for self-efficacy (a) and quality of life (b) at pre-, post-, and four-week follow-up for the three groups from a marginal regression model with generalized estimating equations adjusted for sex and functional class.

QOL scores improved by 3.58 ± 1.10 in the CPR group compared with an improvement of 3.67 ± 0.99 in the CR group (*P* for difference = 0.001). These differences were maintained at follow-up, with no significant change observed between the two groups (*P* for difference = 0.647). The control group showed no significant changes in QOL scores in the pre-, post-, and follow-up tests. QOL declined significantly over time (*P* = 0.002), but the results showed neither differences by group (*P* = 0.787) nor a group-by-time interaction (*P* = 0.066; Figure 4b).

The PA amount increased by 1746.99 ± 108.71 (*P* for difference < 0.001) in the CPR group compared with an increase of 1663.62 ± 175.62 in the CR group (*P* for difference < 0.001). These differences were maintained at follow-up. The control group showed no significant changes in PA amount in the pre-, post-, and follow-up tests. The PA amount increased significantly according to group (*P* = 0.021) and time (*P* < 0.001), showing a significant group-by-time interaction (*P* = 0.008; Table 4). Additionally, the amount of PA increased. The number of patients in the HEPA group significantly increased from 33.3% to 83.3% in the CPR group and from 25.0% to 75.0% in the CR group. At follow-up, the rates were 75.0% in the CPR group and 66.6% in the CR group (Table 4). However, we observed no significant differences between the CPR and CR groups regarding CPET (exercise time, metabolic equivalent, and maximal oxygen uptake) in the pre-, post-, and follow-up tests. The control group also showed no significant changes in the CPET variables in the pre-, post-, and follow-up tests (Table 4).

**Table 4.** Changes in Exercise Capacity of the Three Groups.

|  | CPR group | CR group | Control group | Time | Group | Interaction |
| --- | --- | --- | --- | --- | --- | --- |
|  | Estimated mean<br>(SE),<br>N = 12 | Estimated mean<br>(SE),<br>N = 12 | Estimated mean<br>(SE),<br>N = 12 | P-value |  |  |
| IPAQ |  |  |  |  |  |  |
| Physical activity, (METs-min/week) |  |  |  |  |  |  |
| Pre-test | 711.24 (166.53) | 712.20 (116.30) | 718.12 (142.87) | < 0.001 | 0.021 | 0.002 |
| Post-test | 1746.99 (108.71) | 1663.62 (175.62) | 950.87 (148.31) |  |  |  |
| Follow-up test | 1716.53 (123.29) | 1564.12 (128.16) | 913.53 (134.58) |  |  |  |
| HEPA, n (%) |  |  |  |  |  |  |
| Pre-test | 4 (33.3) | 3 (25.0) | 4 (33.3) | < 0.001 | 0.049 | < 0.001 |
| Post-test | 10 (83.3) | 9 (75.0) | 3 (25.0) |  |  |  |
| Follow-up test | 9 (75.0) | 8 (66.6) | 3 (25.0) |  |  |  |
| CPET |  |  |  |  |  |  |
| Exercise time, (Min) |  |  |  |  |  |  |
| Pre-test | 10.05 (0.53) | 10.18 (0.30) | 10.72 (0.39) | 0.916 | 0.379 | 0.420 |
| Post-test | 10.42 (0.55) | 10.13 (0.48) | 10.60 (0.44) |  |  |  |
| Follow-up test | 10.31 (0.60) | 9.94 (0.43) | 10.77 (0.22) |  |  |  |
| Metabolic equivalent |  |  |  |  |  |  |
| Pre-test | 8.70 (0.47) | 8.90 (0.35) | 9.26 (0.43) | 0.417 | 0.787 | 0.043 |
| Post-test | 9.20 (0.53) | 8.93 (0.31) | 9.04 (0.51) |  |  |  |
| Follow-up test | 9.39 (0.55) | 8.77 (0.32) | 9.22 (0.45) |  |  |  |
| Maximal oxygen uptake (VO<sub>2</sub> max), (mL/Kg/min) |  |  |  |  |  |  |
| Pre-test | 30.44 (1.63) | 29.74 (0.99) | 32.34 (1.50) | 0.065 | 0.530 | 0.141 |
| Post-test | 32.23 (1.84) | 30.53 (1.03) | 31.63 (1.81) |  |  |  |
| Follow-up test | 32.38 (2.11) | 30.60 (1.09) | 3219 (1.58) |  |  |  |

Data are expressed as estimated mean ± standard error or number (percentage).
CPR indicates cardiac and psychosocial rehabilitation; CR, cardiac rehabilitation; IPAQ, international physical activity questionnaire; HEPA, Health enhancing physical activity; and CPET, cardiopulmonary exercise testing.

## DISCUSSION

We examined the effectiveness of an intervention program combining a CBT-based psychological intervention and CR targeting young adults with FC and psychological distress. The eight-week intervention program significantly reduced anxiety and depression scores compared with only CR or usual care. The reduced anxiety and depression scores were maintained at the four-week follow-up. This is partly consistent with the results of Kovacs et al.’s eight-session CBT intervention applied to patients with CHD, which showed decreased depressive symptoms with a medium effect size.^20^

Research on psychological interventions among patients with CHD, including patients with FC, is limited.^22^ Psychological interventions for depression and anxiety may result in a moderate reduction in depression and anxiety and may improve mental health-related QOL in adults with CHD or heart failure, according to 21 RCTs (2,591 participants) that included psychological interventions, including CBT.^58^ A recent meta-analysis of 22 RCTs involving nearly 5,000 patients with coronary artery disease revealed that CBT significantly reduced symptoms of depression and anxiety and improved health-related QOL.^59^ The trials involved a treatment duration of longer than 12 weeks and more than 10 sessions.

Holdgaard et al. reported that depression and anxiety reduced and health-related QOL improved by applying five group sessions.^19^ Nuraeni et al. showed effective depression reduction in a program of more than eight sessions and argued that programs of three to six sessions may be ineffective in reducing depression.^18^ Given that intervention frequency may affect efficacy and efficiency, these factors need to be considered when designing intervention plans.^60^

Individual and group CBT are equally effective,^61^ and adults with CHD frequently seek opportunities to interact with their peers.^62,67^ Moreover, face-to-face and Internet-based CBT have the same effect.^63^ Therefore, Internet-based group CBT may be considered as a method to increase the effectiveness of CBT interventions by increasing the number of sessions and lowering the dropout rate.

Our results showed that the reduced depression and anxiety scores were maintained in the follow-up test one month after the intervention ended. This is consistent with the results of a systematic review and meta-analysis of RCTs in patients with coronary artery disease.^18^ The rate of depression reduction was significantly better in the short-than in the long-term follow-up, which may indicate that CBT rapidly reduces depression. However, nearly all studies have shown that depression lessens over time, perhaps because mild depression resolves on its own^64^ or because of an effect of CR^65^ applied in combination. Future studies should explore the effects of CBT in patients with more severe depressive symptoms to identify CBT’s impact on depression in patients with FC in the short-, medium-, and long-term follow-up.

Our results also showed that self-efficacy increased after the intervention in the CPR group and was maintained at follow-up. This result is consistent with the finding that CBT predicts greater self-efficacy in managing insomnia symptoms.^66^ However, other studies have shown no significant difference in self-efficacy after CBT interventions targeting cardiovascular diseases.^22^

QOL also significantly increased after the intervention but was not significant at the four-week follow-up. These results are consistent with the findings of Kovacs et al., who used the same tool with adults with CHD.^20^ However, contradictory results have been reported in patients with symptomatic paroxysmal atrial fibrillation (AF), in which AF-related QOL improved significantly through online CBT.^67^ We propose to develop a Fontan-specific QOL tool and use it to repeat this study’s tests in future research.

CR interventions can alleviate psychosocial problems in patients with FC. We examined their effect when combined with a CBT-based intervention and found that the self-reported activity level of the group that experienced a combined CBT and CR intervention increased significantly compared with that of the control group and the group that underwent only CR. However, the groups showed no significant differences in exercise capacity (VO2 max, METs, and exercise time). A previous study similarly showed improvement in a CR compliance group in patients with coronary artery disease who received CBT compared with the control group.^19^ However, a systematic literature review that analyzed the results of exercise programs applied to patients with FC under 20 years of age reported a significant enhancement in at least one measure of maximal or submaximal exercise capacity.^68^ Another systematic review that analyzed 16 exercise training studies involving 264 patients with FC with an average age of 8.7–31 years showed improvements in peak oxygen uptake, lung function, PA level, and QOL.^69^ These findings are contrary to our results. CR programs for patients with FC may offer clinical benefits by including exercises that increase muscle mass and strength and employing peripheral pumps to augment the increased physiological response to higher oxygen demand during exercise.^31,41^

Furthermore, to significantly improve VO2 peak or VO2 max in patients with FC, an exercise program should be implemented for three months to one year, with a weekly exercise time of 90–180 minutes.^31^

In our study, the absence of improvement in exercise capacity (VO2 max) may be because we employed an eight-week program focusing on aerobic exercise for a rather small sample. Therefore, future studies should increase the sample size and apply a structured CR program of more than 12 weeks that includes aerobic, resistance, lower extremity strength, and ventilator muscle exercises.^31,41,55^

To the best of our knowledge, this study is the first attempt to apply a CBT-based psychological intervention and CR to adult patients with FC and report positive results. However, this study has several limitations. For example, the generalizability of our results is limited by our use of a single cardiac center, where all participants had their follow-up, and our relatively small sample size. Additionally, our study targeted patients with relatively mild symptoms who had signs of depression or anxiety but did not receive a psychiatric diagnosis or treatment. The CPR group was more homogeneous and thus benefited from group sessions.^19^ However, our results may not apply to patients with FC with moderate-to-severe depressive or anxiety symptoms.

Moreover, we did not measure the negative effects of focusing on psychological problems in the intervention, nor did we determine the effectiveness of CBT treatment in relation to a sham intervention. Patients in the CPR group underwent both CBT and CR; this involved more contact time, which may have contributed to the program’s effectiveness or a belief bias for the patients and healthcare teams. Treatment efficacy tends to be influenced by the therapeutic relationship factors in psychotherapy.^19,70^ Finally, telemonitoring was conducted by a research nurse while CR was being applied. We also considered PA quantities that were self-reported by the patients, which may have been inaccurate. As seen in several recent studies, efficient monitoring and more objective data can be obtained by using mobile apps^71^ and wearable devices such as Fitbits.^72^

### CONCLUSIONS

CPR can be used as part of an intervention to alleviate emotional distress and improve self-efficacy and QOL in young adults with FC. Future efforts are required to increase the efficacy of home-based CR.

## Data Availability

The data will be made available in aggregated form by the authors upon request

## Nonstandard Abbreviations and Acronyms

BDI: Beck Depression Inventory
CBT: Cognitive Behavioral Therapy
CHD: Congenital Heart Disease
CPET: Cardiopulmonary Exercise Testing
CPR: Cardiac and Psychosocial Rehabilitation
CR: Cardiac Rehabilitation
FC: Fontan Circulation
GSE: General Self-Efficacy
HADS: Hospital Anxiety and Depression Scale
HADS-D: Hospital Anxiety and Depression Scale – Depression
HADS-A: Hospital Anxiety and Depression Scale – Anxiety
HEPA: Health-Enhancing Physical Activity
IPAQ: International Physical Activity Questionnaire
LAS: Linear Analog Scale
MET: Metabolic Equivalent
PA: Physical Activity
QOL: Quality of Life
RCT: Randomized Controlled Trials
VO2 max: Maximal Oxygen Uptake

## CLINICAL PERSPECTIVES

### What is new?

Combining cardiac rehabilitation and cognitive behavioral therapy enhances the mental health of adults with Fontan circulation by reducing anxiety and depression. This integrated approach offers a novel method for addressing both physical and psychological challenges and improving overall patient outcomes.

### What are the clinical implications?

Integrating cardiac rehabilitation and psychosocial interventions such as cognitive behavioral therapy into routine care enhances the management of adults with Fontan circulation. This approach addresses both psychological and physical challenges and reduces anxiety and depression while improving quality of life. These findings suggest a shift toward a more comprehensive care model that promotes better long-term outcomes and holistic patient management in clinical practice.

## Acknowledgments

The authors would like to thank all participants and their parent(s) or guardian(s) for their involvement in this study.

## Ethics Approval and Consent to Participate

This study was approved by the Institutional Review Board of the Samsung Medical Center and prospectively registered with the Korean Clinical Trials Registry (KCT 0007737). All procedures were performed in accordance with the relevant guidelines and regulations of the Institutional Review Board. All participants provided written informed consent.

